# Integrating Lung Tissue-based Transcriptome-Wide Association Study with Single-cell RNA-sequencing Uncovers Susceptibility Genes and Cell Types Underlying Lung Cancer Risk

**DOI:** 10.64898/2026.03.19.26348840

**Authors:** Shuai Xu, Jiajun Shi, Bingshan Li, Xiao-Ou Shu, Ran Tao, Hui Cai, Wanqing Wen, Stephen A. Deppen, Xin Maizie Zhou, Lili Xu, Jifeng Wang, Jie Wu, Yaohua Yang, Xingyi Guo, Wei Zheng, Jirong Long, Qiuyin Cai

## Abstract

Genome-wide association studies (GWASs) have identified approximately 100 loci for lung cancer, but potential causal genes remain largely unknown. To address this, we conducted a lung tissue-specific transcriptome-wide association study (TWAS). Gene expression prediction models were constructed using data of adjacent normal lung tissues from our Vanderbilt Thoracic Biorepository (N=314) and normal lung tissues from the GTEx (N=466) and then applied to our lung cancer GWAS meta-analysis (55,174 cases and 1,294,174 controls). We identified 109 unique risk genes for lung cancer and its histological subtypes. Of them, 71 unique genes were novel discoveries, and 13 unique genes reside in novel loci. Smoking-conditional analysis revealed that 52 unique genes are unrelated to smoking behavior. Seven unique genes showed cell-type-specific colocalization within potential risk cell types, including the alveolar type I and II, dendritic, and natural killer cells. Seventeen unique genes are targeted of 58 drugs that have been approved or in Phase II or III trials. In addition, 22 unique potential causal genes were supported by both Mendelian randomization and colocalization. Functional validation identified three genes through *in vitro* knockdown experiments. Our study identified new lung cancer candidate risk genes and offered insights into lung cancer biology and future translational utilities.

## Introduction

Lung cancer is one of the most common cancers and the leading cause of cancer-related death worldwide for both men and women.^1,2^ While cigarettes smoking is the primary risk factor, a twin study suggested that genetic factors contribute moderately (>20%) to lung cancer susceptibility.^3^ Genome-wide association studies (GWASs) have identified about 100 risk loci for lung cancer,^4–6^ however, most are located in non-coding regions, making it challenging to pinpoint potential causal genes at these loci.

Transcriptome-wide association studies (TWASs) have been developed to address this gap by linking expression levels predicted by genetic variants to disease risk and have been shown to be a powerful approach to identifying candidate causal genes.^7^ By leveraging lung tissue-specific TWAS, we can identify susceptibility genes that are unique to the target organ, providing a more biologically relevant perspective on lung cancer etiology. To date, lung-tissue-based TWASs have identified nearly 100 candidate genes for the risk of lung cancer across diverse populations.^8–13^ But these studies had modest statistical power due to moderate sample sizes of the GWAS and/or limited datasets for gene expression prediction. Availability of multi-omics resources and multiple gene expression reference panels combined with multi-tissue TWAS algorithms offers opportunities to improve power for gene discovery.^14^ In addition, identifying susceptibility genes independent of smoking behaviors could help understand lung cancer etiology. Furthermore, understanding the cellular context in which TWAS-implicated genes influence disease risk remains a critical unmet need, especially within epithelial cells where lung cancer commonly originates. Emerging methods can integrate TWAS gene sets with single-cell RNA sequencing (scRNA-seq) data to highlight potential disease-relevant cell types and refine biological interpretation.^15^

In this study, we performed the largest-ever TWAS to identify genes associated with risk of lung cancer and its major histological subtypes in European-ancestry populations by using our treatment-naïve, tumor-adjacent normal lung tissue samples from the Vanderbilt Thoracic Biorepository (VTB), normal lung tissue samples from the Genotype-Tissue Expression project (GTEx), and our newly generated meta-analysis results combining four publicly available lung cancer GWAS data with a total of 55,174 cases and 1,294,174 controls. We then explored putative smoking-unrelated risk genes to help understand the potential lung cancer etiology other than cigarette smoking. We also integrated our findings with scRNA-seq data from the Human Lung Cell Atlas^16^ to explore in which cell types these genes may affect lung cancer risk. Furthermore, Bayesian colocalization, Mendelian randomization (MR), *in vitro* experiments, scRNA-seq analysis, cell type-specific expression quantitative trait loci (ct-eQTL) colocalization, and drug target queries provided functional context and translational relevance. Together, these efforts aim to clarify the genetic architecture and cellular mechanisms of lung carcinogenesis, informing precision prevention and therapeutic target discovery.

## Results

### Prediction model building

The analytical workflow and design of this study are illustrated in Figure 1. We generated and used transcriptomic and genetic data from our VTB study, along with publicly available GTEx data to predict gene expression levels. In this study, we focused on protein-coding genes and long noncoding RNAs, collectively referred to as genes for simplicity. Among 21,166 highly expressed autosomal genes, expression levels of 14,257 (67.4%) genes were predicted by using their *cis* variants in the VTB study. Of them, 8,492 (59.6%) genes achieved prediction performance of *R* > 0.1 and *P* < 0.05. *R* > 0.1 represents correlation of >10% between the observed and the predicted expression levels. On average, a median of 24 (interquartile ranges [IQR]: 15-36) genetic variants were used to build each model. For GTEx data, we successfully built genetic imputation models for 19,010 (72.0%) out of 26,418 highly expressed autosomal genes. A total of 10,661 (56.1%) reliably predictive models were constructed with a median of 22 (IQR: 14-34) genetic variants used per model. Combining the data from VTB and GTEx data, prediction models were successfully built for 12,902 unique genes. Among them, prediction models were successfully built in both VTB and GTEx datasets for 6,251 genes, with a correlation in their prediction performance of 0.63 (*P* < 0.001) across the two sets of models.

**Figure 1.**
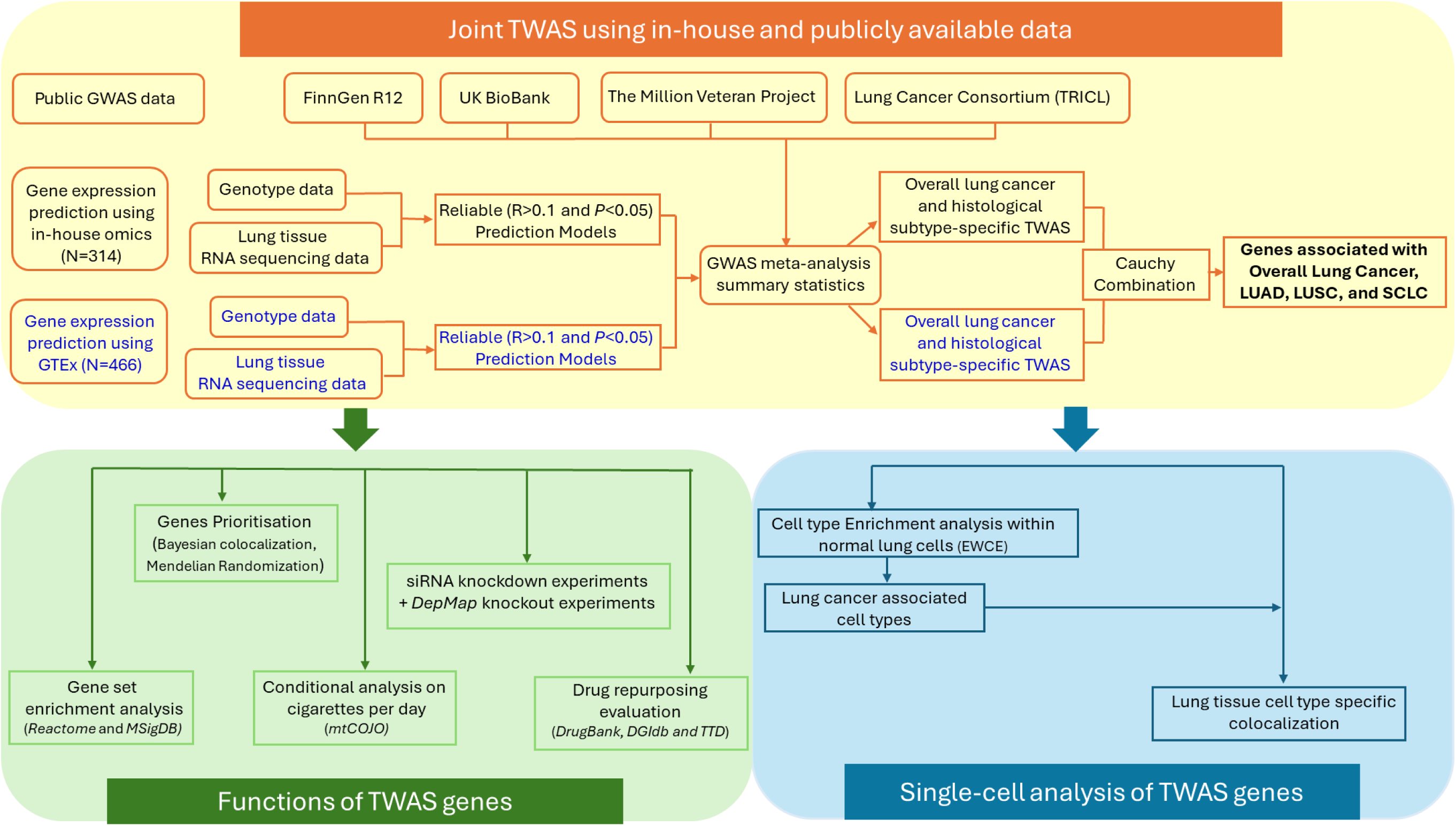
Analytic flowchart of study design.

### Identifying putative risk genes for overall lung cancer

We first applied genetic prediction models derived from VTB and GTEx with lung cancer GWAS data under S-PrediXcan framework and then combined *P* values of TWAS results from those two sets of models using the aggregated Cauchy association test (ACAT). The ACAT combines only *P* values but not Z scores. We observed that Z scores across VTB and GTEx were moderately consistent, with a correlations of 0.73 (*P* <0.001), confirming concordant effect directions and ACAT’s applicability. Of 12,902 tested genes, genetically predicted expression levels of 87 genes were significantly associated with overall lung cancer risk after Bonferroni correction at *P* < 3.87×10^-6^ (**Figure 2A** and **Table S2**). Of the 87 significant TWAS-identified genes, 56 (64.4%) have not been reported in any previous lung cancer TWAS. Eight genes in five loci, including *ENSG00000261189* (6p24.3), *ENSG00000285668* (17q21.31), *MAPT* (17q21.31), *MAPT-IT1* (17q21.31), *LRRC37A2* (17q21.31), *ENSG00000277476* (17q24.2), *LPAR2* (19p13.11), and *RPS5* (19q13.43), were located more than 2Mb away from any of the risk variants identified in lung cancer GWAS conducted previously or in our recent meta-analysis, representing novel risk loci. Of 59 previously reported genes across 11 loci (**Table S3**),^8,9^ we found 30 (50.8%) genes across nine loci at *P* < 1×10^-4^ in our study, supporting the validity of our approach.

**Figure 2.**
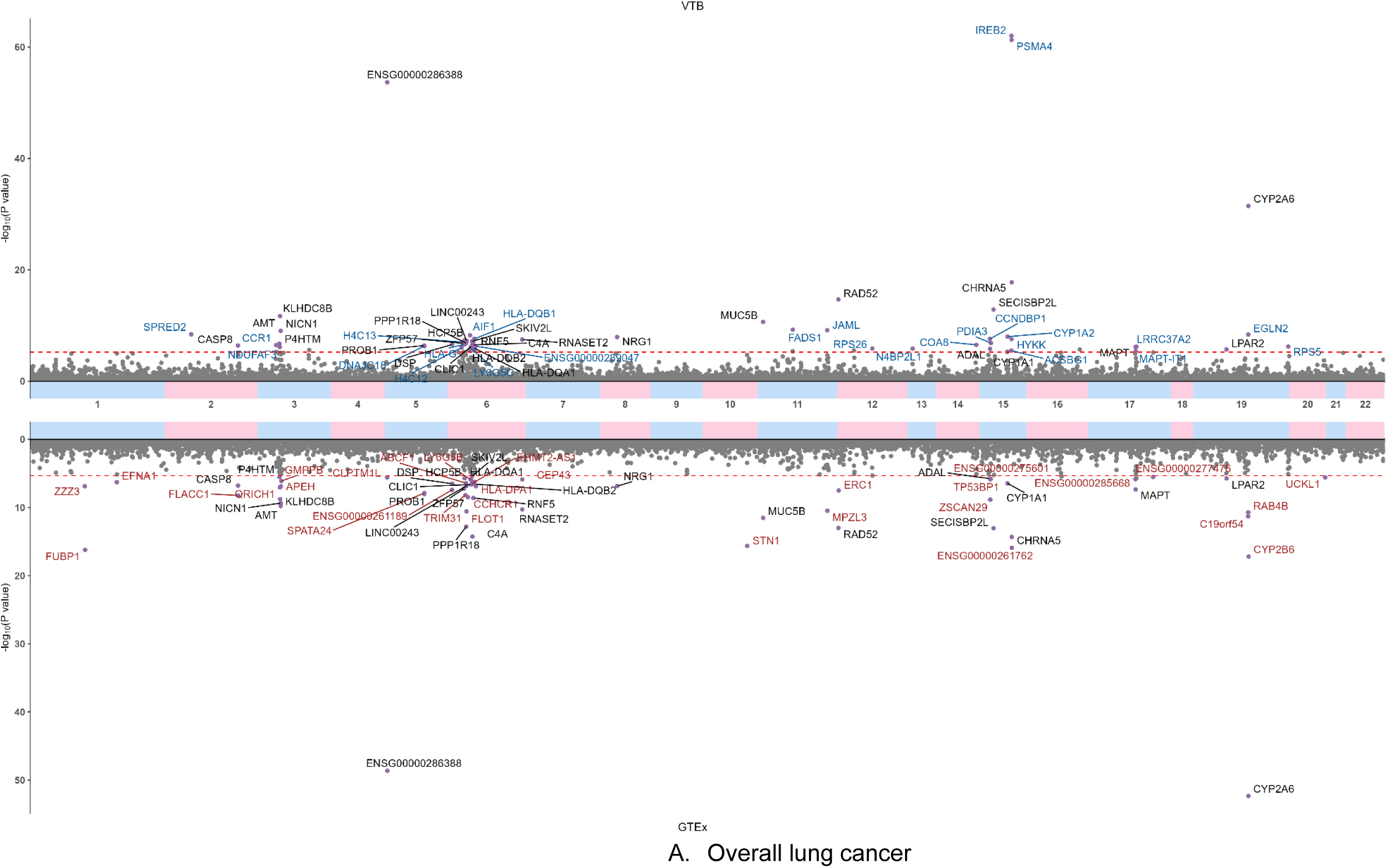
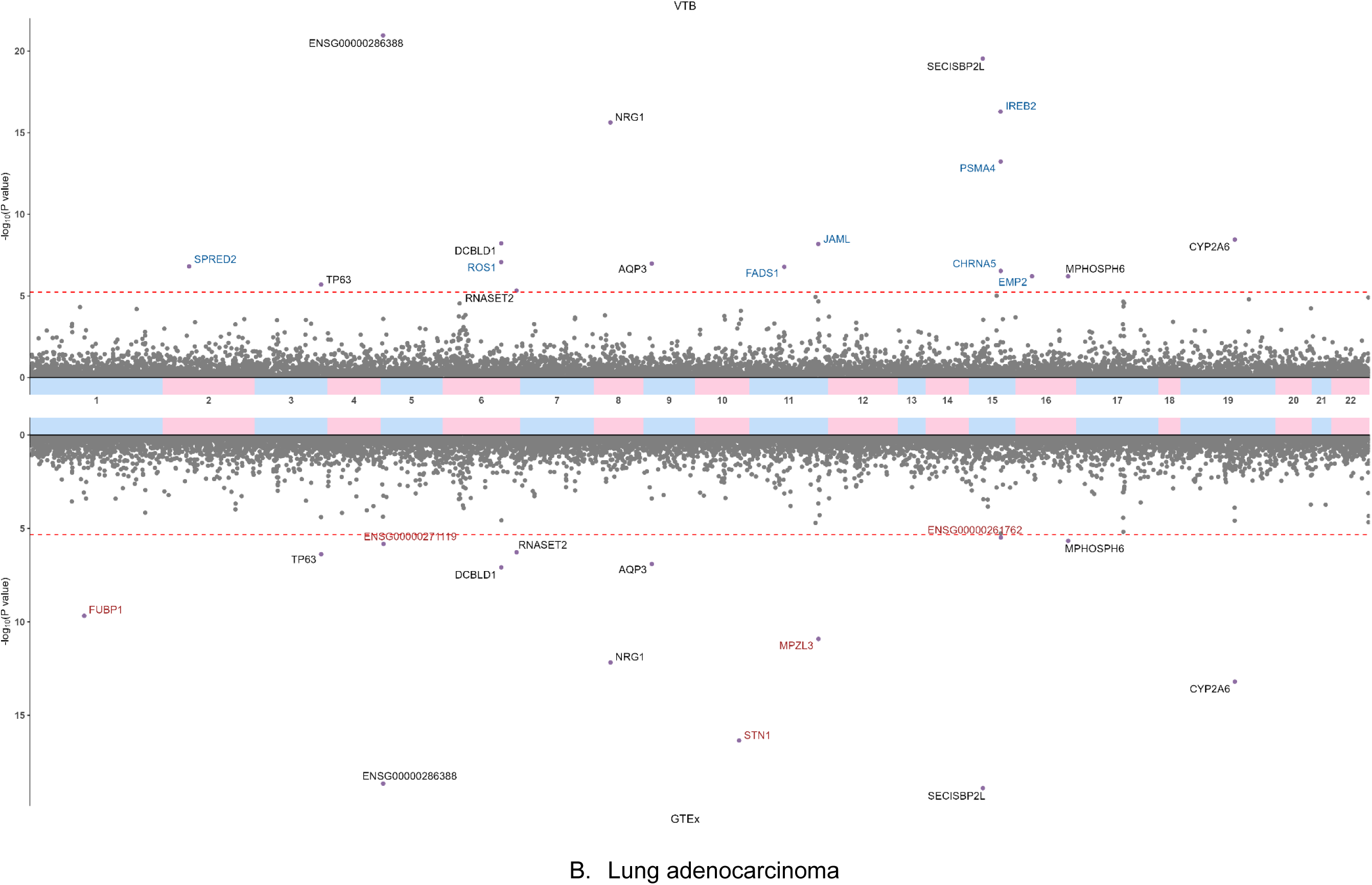
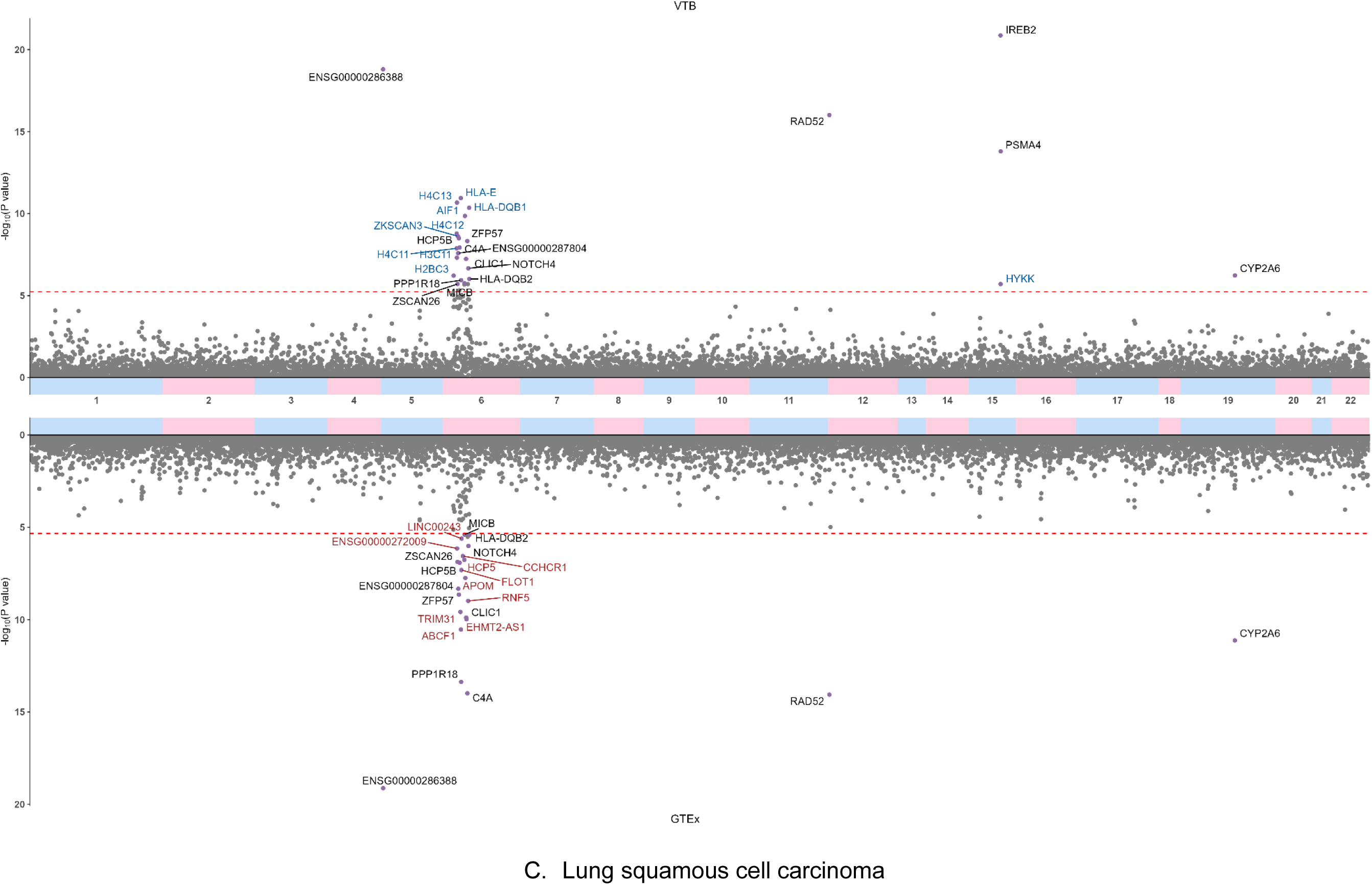
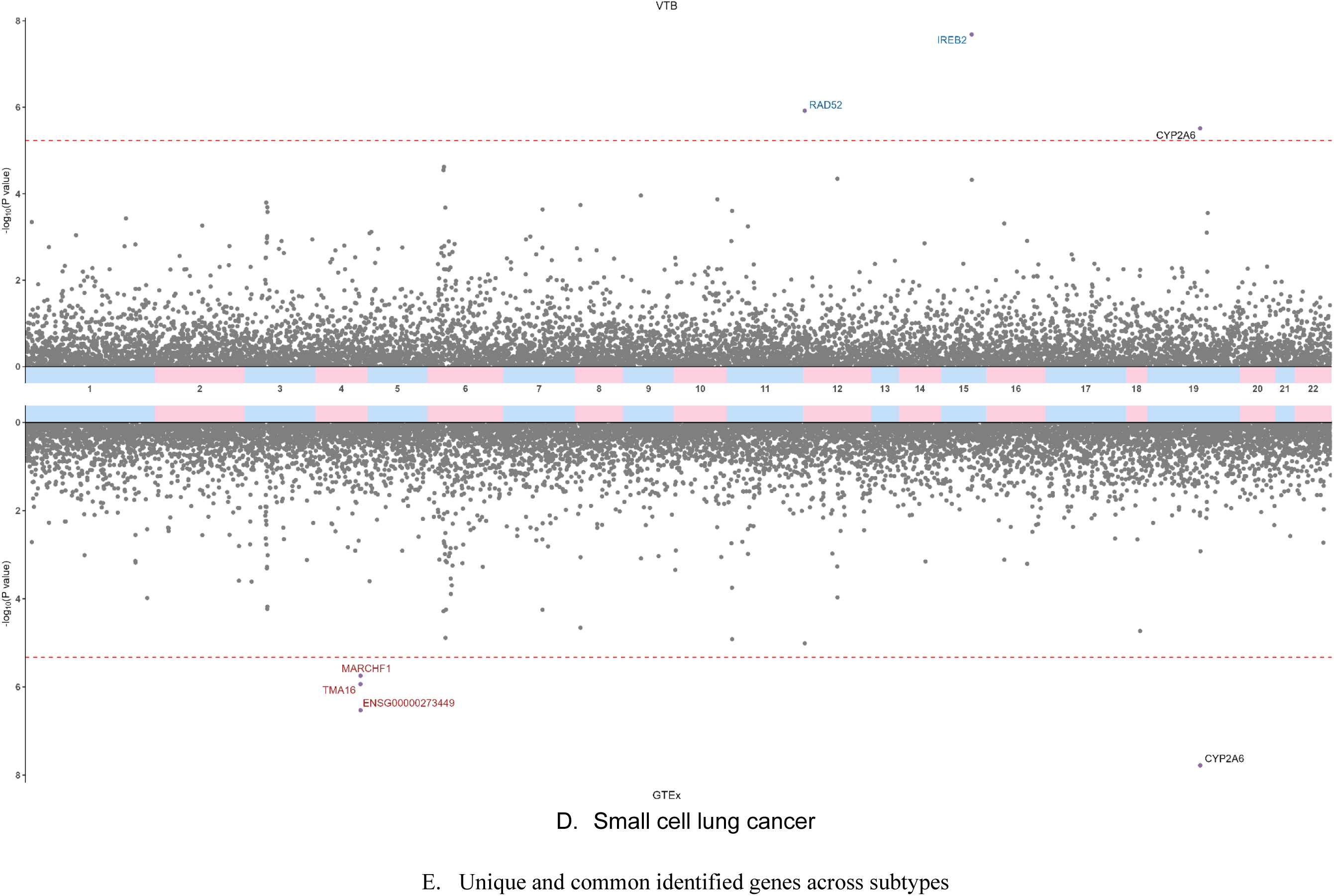
TWAS identified putative genes for the risk of overall lung cancer (A), adenocarcinoma (B), squamous cell carcinoma (C), and small cell lung cancer (D). The upper plots are TWAS results by integrating prediction models derived from 314 VTB participants, and the lower ones are TWAS results by integrating prediction models constructed from 466 GTEx participants. Genes identified by both studies are highlighted in black. Genes identified only by VTB are highlighted in blue. Genes identified only by GTEx are highlighted in red.

### Identifying risk genes for lung cancer histological subtypes

The genetic etiology of lung cancer may vary significantly across its histological subtypes. To further identify putative risk genes specific to lung cancer major histological subtypes, we conducted TWASs that followed the same pipeline for lung adenocarcinoma (LUAD), squamous cell carcinoma (LUSC), and small cell lung cancer (SCLC), respectively. At the Bonferroni-correction significance level of *P* < 3.87×10^-6^, we identified genetically proxied expression levels of 22 genes associated with LUAD risk (**Figure 2B** and **Table S2**), 35 genes with LUSC risk (**Figure 2C** and **Table S2**), and six genes with SCLC risk (**Figure 2D** and **Table S2**). Among histological subtypes associated risk genes, seven (*TP63, ROS1, ENSG00000271119*, *DCBLD1, AQP3,* and *EMP2*) were specific to LUAD risk; 12 (*H2BC3, H4C11, H3C11, ZSCAN26, ZKSCAN3, ENSG00000287804, HLA-E, MICB, ENSG00000285761, HCP5, APOM,* and *NOTCH4*) were exclusively associated with LUSC risk; and three (*TMA16*, *MARCHF1*, and *ENSG00000273449*) were exclusively associated with SCLC risk.

Nine LUAD-, 14 LUSC-, and two SCLC- associated risk genes have not been reported in any previous lung cancer subtype-specific TWAS studies. Additionally, two LUAD- (*RNASET2* at 6q27 and *EMP2* at 16p13.13) and three SCLC- (*TMA16* and *MARCHF1* at 4q32.3 and *ENSG00000273449* at 4q32.2) associated risk genes were located at least 2Mb away from histological subtypes-specific GWAS index variants identified previously or by our meta-analysis, indicating potential novel risk loci. To replicate the previously reported histological subtype-specific TWAS signals, we compared our results with previous European-ancestry lung tissue TWASs, which identified 20 LUAD genes across 11 loci, 50 LUSC genes across seven loci, and nine SCLC genes across four loci. We replicated 14 (70.0%) LUAD genes across ten loci, 27 (54.0%) LUSC genes across six loci, and five (55.6%) SCLC genes across two loci at threshold *P* < 1×10^-4^ (**Table S3**).

In total, we identified 109 unique susceptibility genes for overall lung cancer or major histology subtypes, including 71 unique novel susceptibility genes and 13 unique genes mapping to new risk loci. Gene-set enrichment analysis via *enrichR* webtool^9,17^ found that 97 protein-coding genes were enriched in 28 *Reactome* pathways, most of which are immune-related, including the PD-1 signaling and the interferon gamma signaling pathways (false discovery rate [FDR] < 5%) (**Figure S1** and **Table S5**).

### Conditioning analysis on cigarette smoking

Cigarette smoking is a major risk factor of lung cancer. To identify lung cancer risk genes independent of smoking behavior, we performed a multi-trait conditional and joint analysis (mtCOJO)^22^ on our newly generated lung cancer GWAS, conditioning on a GWAS of cigarettes per day^23^—this smoking trait showing the strongest genetic correlation with lung cancer.^6^ We then conducted TWAS using the lung cancer risk GWAS summary statistics results after adjusted for the smoking GWAS. After adjusting for cigarettes per day, 30 (34.5%) overall lung cancer-associated genes remained at the previous threshold (*P* < 3.87×10^-6^) (**Table S9**). As expected, well-known smoking-related genes like *CHRNA5* and *CYP2A6* were no longer significant, validating this approach. In histological-subtype analyses, 14 (63.6%) LUAD-, 20 (57.1%) LUSC-, and one (16.7%) SCLC- associated genes remained significant after adjusting for smoking (**Table S9**). Of note, associations of five genes with overall lung cancer risk became more significant and reached the significance threshold after adjustment, namely *CLDN18* (*P* = 4.15×10^-6^ to 9.92×10^-7^), *PBK* (*P* = 9.30×10^-6^ to 9.16×10^-7^), *BHLHE41* (*P* = 5.66×10^-6^ to 1.33×10^-6^), *SLC12A9* (*P* = 1.96×10^-4^ to 3.60×10^-6^), and *ENSG00000256234* (*P* = 1.65×10^-5^ to 2.22×10^-6^). Additionally, the associations of *HLA-DQA1* with LUSC risk became more significant (*P* = 1.19×10^-5^ to 4.30×10^-9^) after conditioning. Overall, 57 unique risk genes are unrelated to smoking behavior, including 52 TWAS-identified and five additional unique genes through smoking-conditional analysis. These smoking-independent risk genes may help enhance the understanding of lung carcinogenesis other than cigarette smoking.

### Drug targets query

To assess whether the implicated genes may serve as potential therapeutic targets, we queried three major drug databases, namely, DrugBank,^24^ ChEMBL,^25^ and the Therapeutic Target Database (TTD).^26^ These resources were used to identify known drug-gene relationships for the 97 unique TWAS-associated protein-coding genes. From these databases, 17 genes encoding proteins showed targetability by 58 distinct drugs that are either FDA-approved (n = 37) for clinical use or are currently investigated in phase II or III (n = 21) clinical trials (**Figure 3** and **Table S6**). Of note, eight FDA-approved drugs targeting the proto-oncogene tyrosine-protein kinase ROS1, including Crizotinib, Entrectinib, Lorlatinib, Repotrectinib, and Taletrectinib, have been used for the treatment of non-small cell lung cancer (NSCLC)^27^. In addition, Daprodustat, which inhibits Egl-9 family hypoxia inducible factor 2 (EGLN2), is an approved drug for the management of anemia.^28^

**Figure 3.**
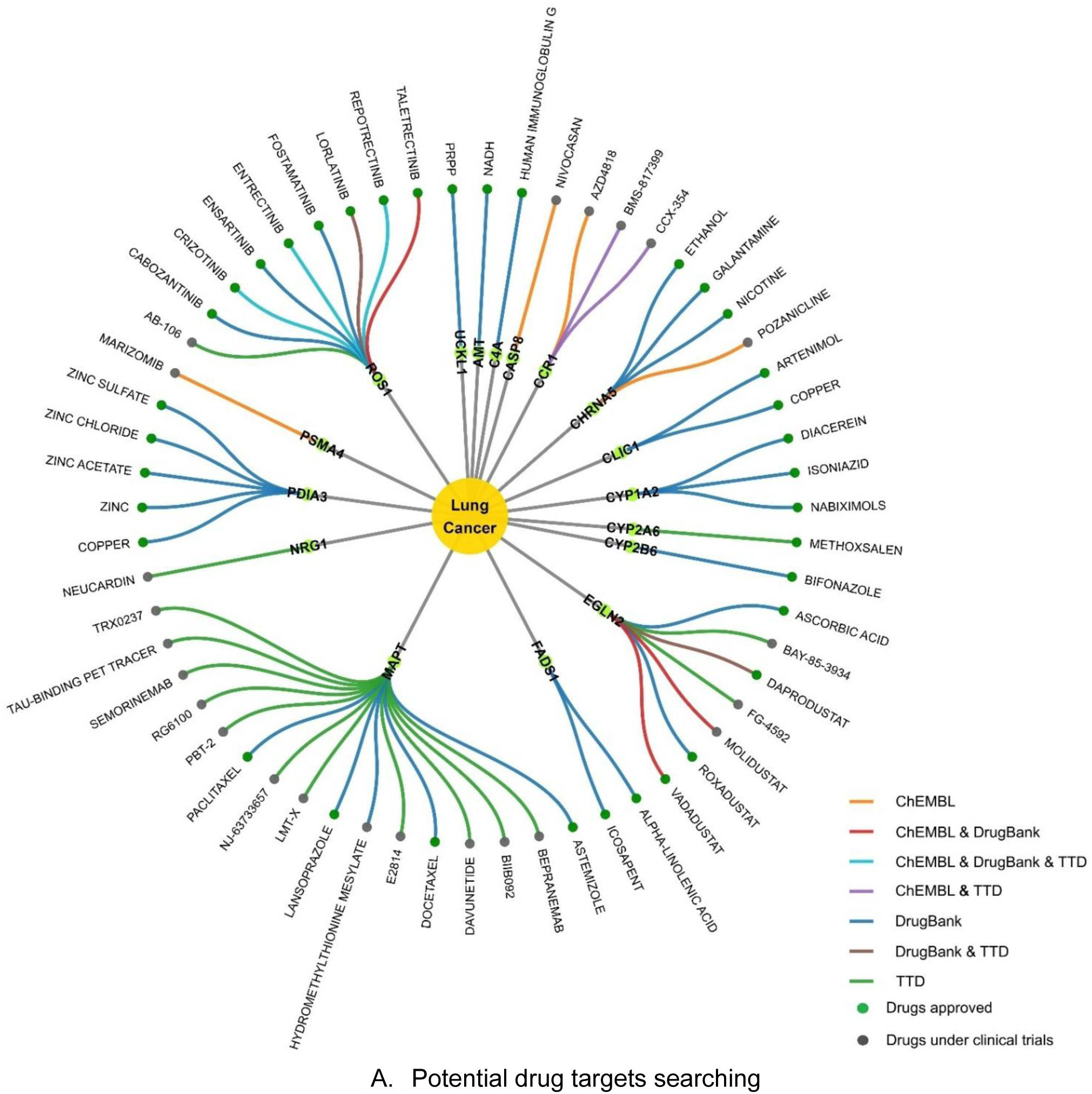
Circular plot of druggable genes and their associated drugs for lung cancer prevention and treatment.

### Discovering enriched cell types and cell type specificity of TWAS-identified genes

Disentangling the cell-type-specific effects of gene expression in lung cancer risk cell types among lung tissues could offer new biological insights. We first adopted the expression weighted cell type enrichment (EWCE) analysis^15^ to discover lung cell types in which TWAS-identified genes were enriched by leveraging scRNA-seq data from normal lung tissue samples from 63 self-reported Europeans generated in the Human Lung Cell Atlas.^16^ We hypothesized that enriched cell types of TWAS-identified genes may serve as candidate risk cell types of developing lung cancer. TWAS genes for overall lung cancer risk were enriched in AT1 cells and dendritic cells (DCs) at *P* < 0.05 (**Figure 4** and **Table S7**). LUAD-associated genes were enriched in AT2 cells, while LUSC-associated genes were enriched in DCs and natural killer (NK) cells at *P* < 0.05. No cell type enrichment reached FDR < 5%. We did not evaluate cell-type-specific enrichment for SCLC due to its insufficient gene number (n = 6). These findings align with established biology: AT1 and AT2 cells are known cells of origin for LUAD,^29,30^ and cigarette smoking, a major risk factor for LUSC, induce the accumulation of lung DCs in a mouse model.^49^

**Figure 4.**
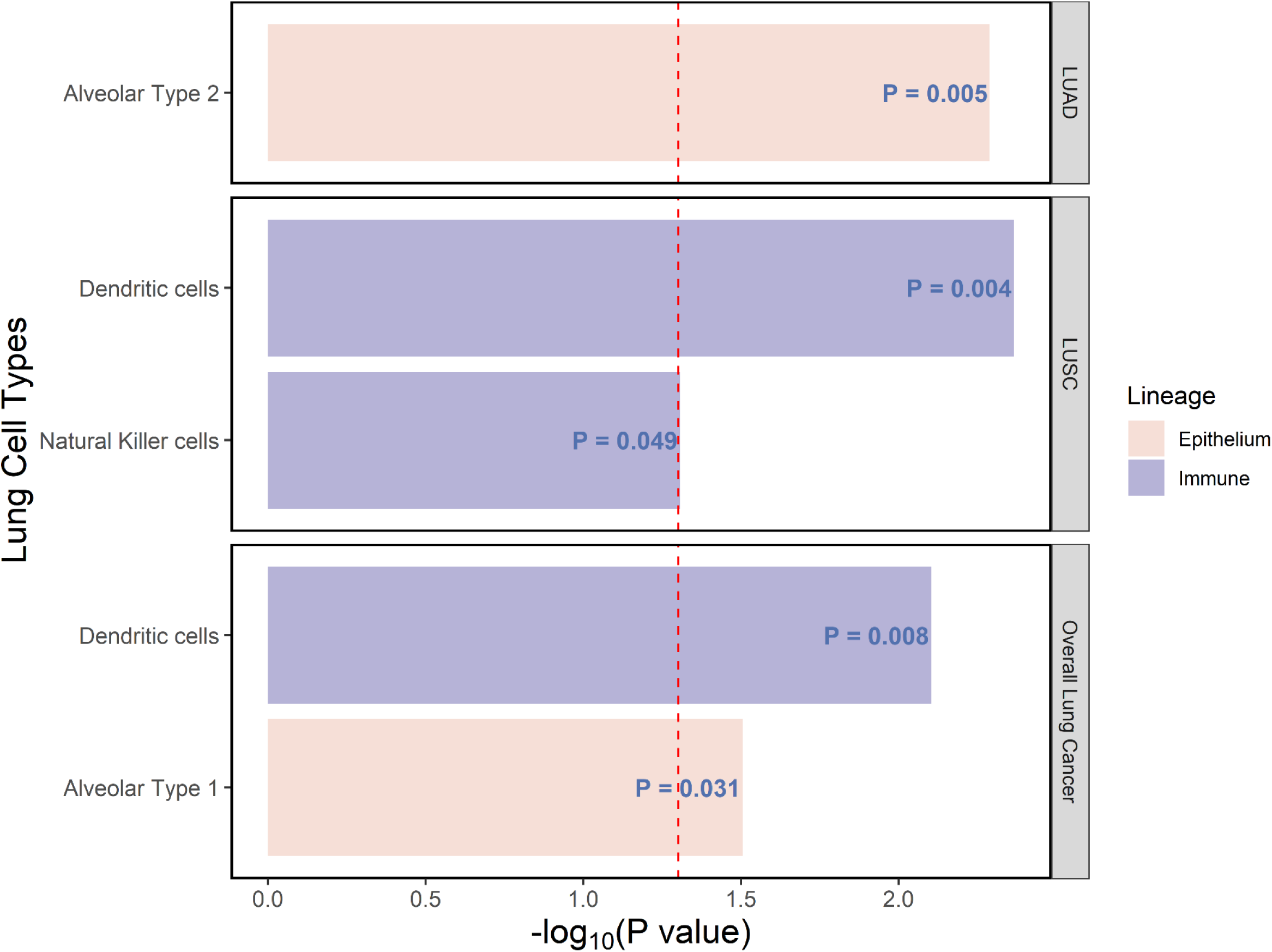
Enriched cell types by histological types of TWAS-identified genes.

We further explored cell-type-specific associations by testing for colocalization between cell-type-specific eQTLs (ct-eQTLs) and lung cancer GWAS in the identified cell types (AT1, AT2, NK, conventional DC types 1 and 2, plasmacytoid DC, and monocyte-derived DC). Six overall lung cancer-associated risk genes, *DSP*, *RPS26*, *FUBP1, RNASET2, RPS5,* and *SECISBP2L,* showed evidence of colocalization in at least one cell type at PP.H4 > 0.70 (**Table S8**). Four LUAD associated risk genes, namely *AQP3*, *FUBP1, SECISBP2L,* and *RNASET2*, demonstrated evidence of colocalization only in AT2 cells (**Table S8**). No colocalization was detected for LUSC associated risk genes in DCs or NK cells, likely attributable to limited ct-eQTL sample sizes (N = 44–59).

### Potential causal genes assessment

To identify likely causal genes among TWAS-identified candidates, we performed Bayesian colocalization^19^ and Summary-data-based MR (SMR)^20^ between expression quantitative trait loci (eQTL) and lung cancer GWAS. We used meta-analyzed eQTLs across VTB and GTEx for these analyses. Of 87 overall lung cancer-associated genes, 15 were supported by both colocalization and SMR algorithms, including *KLHDC8B, AMT, NICN1, DSP, NRG1, ENSG00000261189, STN1, MUC5B, FADS1, COA8, ZSCAN29, SECISBP2L, ENSG00000277476, LPAR2,* and *RPS5* (**Figure 5 and Table S4**). In addition, seven LUAD- (*ROS1, NRG1, AQP3, RNASET2, MPHOSPH6, SECISBP2L,* and *STN1*), one LUSC-(*PPP1R18*), and two SCLC- (*TMA16* and *CYP2A6*) associated risk genes were considered as high-confidence causal genes in both approaches (**Figure 5 and Table S4**). Notably, our identification of two well-known lung cancer oncogenes *NRG1* and *ROS1* demonstrates the validity of our analysis.^21^

**Figure 5.**
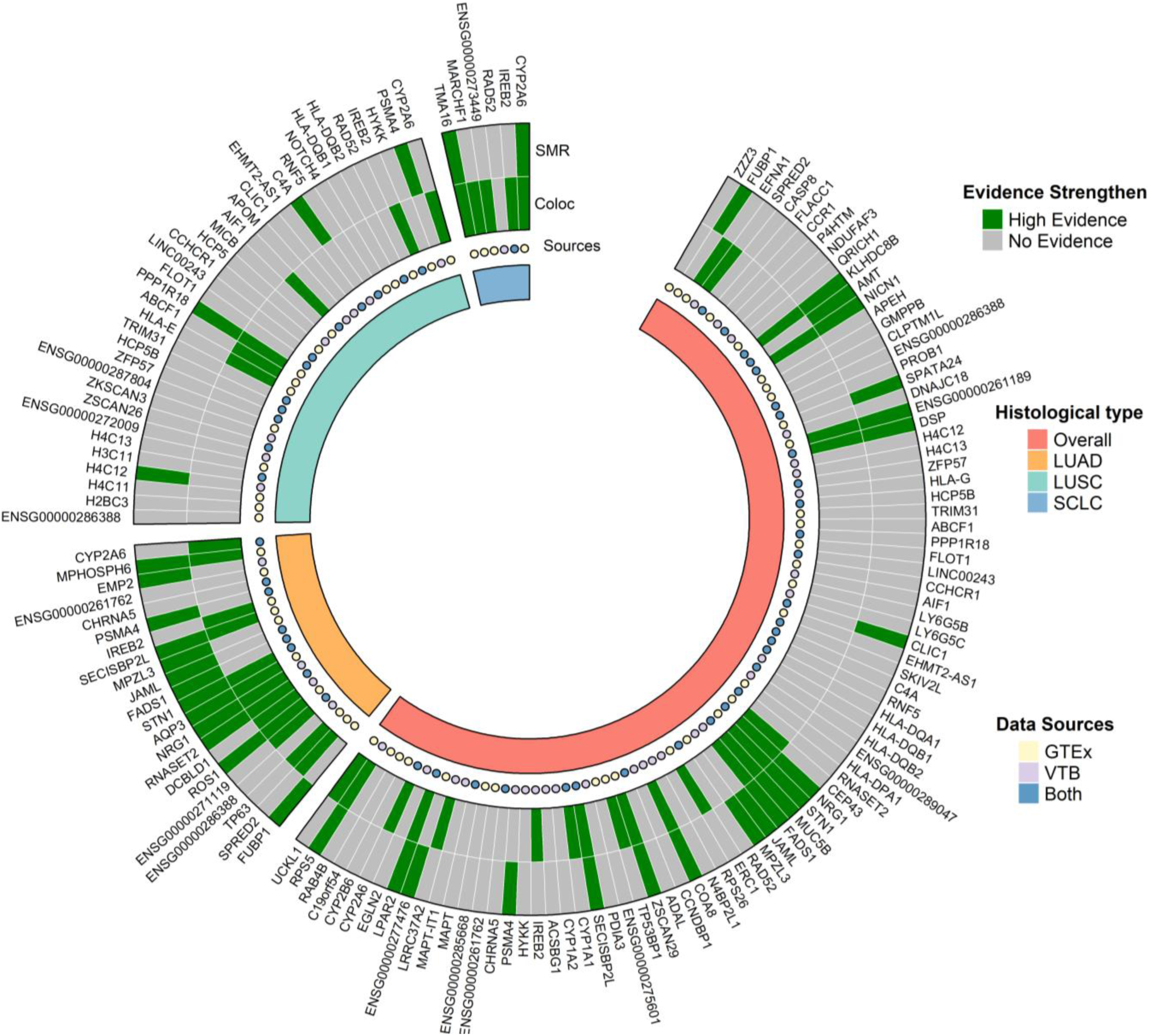
High-confidence putative risk genes for the risk of overall lung cancer, adenocarcinoma, squamous cell carcinoma, and small cell lung cancer supported by Bayesian Colocalization and Summary-data-based Mendelian Randomization. A posterior probability of hypothesis 4 of causal variants shared by both traits > 0.7 was defined as high-confidence risk genes in Bayesian colocalization. Bonferroni corrected *P* < 0.05 and nominal *P* of the heterogeneity in dependent instruments test > 0.05 were considered as high-confidence causal genes.

### Functional assays for putative susceptibility genes

Beyond *in silico* evidence, we further functionally investigated the roles of lung cancer putative risk protein-coding genes in lung cancer cell lines through *in vitro* experiments. We first evaluated gene essentiality of proliferation using the *Chronos* score from CRISPR-Cas9 screen experiments in 126 lung cancer cell lines from the Cancer Dependency Map project (DepMap).^18^ Out of 87 protein-coding genes with available *Chronos* data, five genes (*PSMA4*, *RPS5*, *GMPPB*, *ABCF1*, and *TMA16*) had a median score of < -0.5, suggesting their essential roles in the lung tumor cell proliferation (**Figure S2**).

We then performed small interfering RNA knockdown experiments in two lung cancer cell lines, namely A549 and PC-9. *DNAJC18*, *PSMA4* and *AQP3* genes were selected due to their higher expression levels in both A549 and PC-9 lung cancer cell lines and lack of prior functional characterization. Their genetically regulated expression levels were positively associated with lung cancer risk. Our experiments showed that cell proliferation, colony formation, migration, and invasion decreased after knockdown expressions of the above three genes compared to negative controls at nominal *P* < 0.05 (**Figure 6**). Our *in vitro* experiments demonstrated the potential oncogenic functions of *DNAJC18, PSMA4* and *AQP3* genes, aligning with our TWAS analysis.

**Figure 6.**
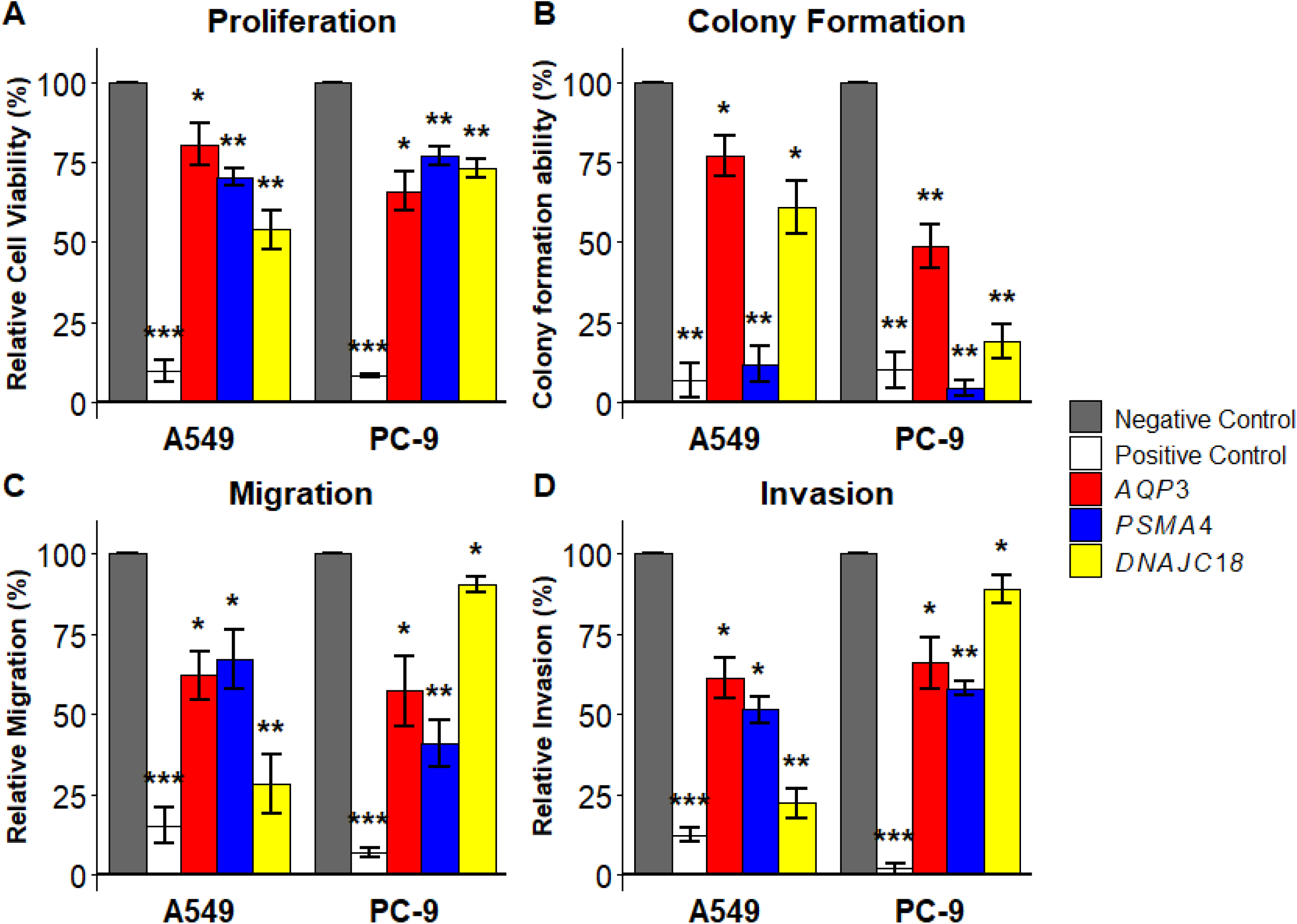
*In vitro* functional assays of *AQP3*, *PSMA4* and *DNAJC18* genes. Knocking down expression of these genes significantly reduced proliferation (A), colony formation (B), migration (C), and invasion (D) in two lung cancer cell lines-A549 and PC-9. Proliferation, colony formation, migration, and invasion were significantly reduced following *AQP3*, *PSMA4* and *DNAJC18* genes knockdown, when compared to the negative control (NC). Statistical significance is denoted by the number of asterisks; * = p < 0.05, ** = p < 0.01, and *** = p < 0.001

## Discussion

In this research, we generated and used transcriptome and genetic data of pathologically confirmed normal lung tissues from 314 VTB primary lung cancer patients, alongside publicly available multi-omics data from 466 GTEx participants for TWAS. We identified 109 unique genes associated with lung cancer susceptibility. Of these, 71 unique genes have not been reported, and 13 unique were located >2Mb away from GWAS index variants. Among these, we uncovered 52 unique smoking-independent candidate genes, and 22 unique candidate causal genes were supported by both colocalization and SMR. *In vitro* knockdown experiments provided additional functional evidence for 3 unique risk genes. Integrative analyses with scRNA-seq data revealed that seven unique risk genes exhibited evidence of colocalization in lung cancer risk cell types, including AT1, AT2, DC, and NK cells. The increased number of candidate susceptibility genes, the identified lung cancer-associated cell types, and cell-type-specific roles of risk genes improve our understanding of the genetic and cellular mechanisms of lung carcinogenesis.

We built genetic prediction models within two large-scale lung-tissue-based RNA-seq eQTL studies: 314 VTB and 466 publicly available GTEx participants. By leveraging both datasets, we substantially expanded our genetic prediction models. The VTB study alone generated genetic prediction models with satisfied performance for 8,492 genes, while GTEx yielded prediction models for an additional nearly 52.0% (n=4,410) genes.

Due to the large sample size of GWAS data and prediction model building data sets, we had higher statistical power than previous studies, which enabled the identification of 71 unique novel genes and replicated previously identified TWAS risk genes. Potential roles of several novel TWAS-identified genes are supported by previous studies. For example, knockdown of the *CCR1* gene significantly reduced the invasiveness of human NSCLC clones, independently of their proliferation.^32^ Desmoplakin (DSP) has been shown to suppress NSCLC, possibly by inhibiting the Wnt/β-catenin signaling pathway in cell lines^33^. As a member of the mucin family, *MUC5B* plays a critical protective role in respiratory airway defense.^34^ Abnormal expression of *MUC5B* was related to NSCLC progression, survival and prognosis.^35,36^ Decreased expression of *FADS1* in tumor tissues of NSCLC patients was associated with worse survival.^37^ *NOTCH4* is a receptor of Notch signaling, which regulates homeostasis and development in multiple tissues.^38^ *NOTCH4* promotes tumor progression of NSCLC, serving as a potential drug target.^39,40^ Loss-of-function mutations in Notch family members are common in LUSC.^41^ Cytochrome P450 (CYP) enzymes CYP1A1, CYP1A2, and CYP2A6 are involved in the metabolic activation of environmental carcinogens such as tobacco-related N-nitrosamines.^42^ Low expression of *ACSBG1* was associated with poor prognosis of lung cancer.^43^ The eQTL of *MPHOSPH6* in GTEx lung tissue colocalized with LUAD risk, leukocyte telomere length, and pulmonary functions.^44^ *ROS1* is an oncogene and a druggable target for NSCLC by several FDR-approved tyrosine kinase inhibitors.^27,45^ *AIF1* correlated with NSCLC prognosis and promoted cell proliferation and migration in A549 cell lines.^46^ Further research is warranted to investigate the underlying biological mechanisms of identified genes in lung cancer development.

In addition to covariate adjustment during genetic prediction model building, we also conditioned GWAS summary-statistic on smoking intensity to attenuate smoking-correlated genetic signals, with a goal of exploring smoking-unrelated susceptibility genes and informing potential lung cancer biology other than cigarette smoking and understanding lung cancer etiology in the understudied never-smoker population. TWAS is underpowered among never-smokers, as the GWAS sample size is small. Only three genes were found to be associated with lung cancer risk among never smokers.^8^ Our smoking-adjusted analysis identified substantially more genes: 44% of the 87 lung cancer-associated genes remained significant, plus five additional genes showed strengthened associations after conditioning on smoking intensity. Of these additionally identified smoking-independent genes, several have been supported by previous studies. Nuclear transcriptional repressor *BHLHE41* was downregulated in tumor tissues and related to poor prognosis of LUAD.^47^ *PBK* was upregulated in lung tumor tissues and negatively associated with survival among lung cancer patients.^48^ These findings suggest potential roles for these genes in lung carcinogenesis beyond smoking-related mechanisms. Larger lung cancer GWAS in never-smokers and sufficiently powered TWAS are needed to validate our results.

To our knowledge, this is the first study that integrates TWAS genes into lung-tissue-based scRNA-seq data to investigate potential genetics and cellular mechanisms of lung cancer development at single-cell resolution. Traditional bulk RNA-seq reflects weighted averages across different cell types, hindering from inferences of cell type specific regulation. However, our bulk TWAS-identified genes were indeed enriched in two epithelial cells of origin and two immune cells, demonstrating the cell-type-specific patterns. The discovery of immune cells also aligns with our pathway enrichment analysis. Our results suggest a potential epithelium-immune crosstalk during lung carcinogenesis. We hypothesized that TWAS-identified genes in immune cells might impact the immune microenvironment and then contribute to lung cancer development among smokers. Natural killer cells play well-established roles in detection and elimination of abnormal cells, preventing tumor development.^50^ Cell-type-specific colocalization analysis supported that 7 unique identified genes play roles in lung carcinogenesis among four potentially affected cell types, providing valuable evidence for lung cancer biology at single cell levels. Future studies are warranted to explore the underlying biological mechanisms and genes in these cell types for their contributions to lung cancer development.

We linked our findings to translational impacts by drug repurposing analysis. From three main drug databases, we identified 17 genes linked to 58 approved or investigated drugs that have been or may be used for lung cancer treatment. For example, eight drugs targeting ROS1 are approved for lung cancer treatment. Lansoprazole, which can bind to MAPT, combined with the use of Gefitinib, showed antitumor effect on A549 cells and mouse xenograft models.^51^ Future trials and studies are needed to validate the repurposability, effectiveness, and efficiency of these drugs for lung cancer prevention and treatment in real-world settings.

There are several limitations of this study. First, lack of normal lung tissue from non-European individuals restricted our study to the European ancestry population, potentially limiting generality. For example, out of eight previously reported TWAS-identified risk genes for lung cancer among Chinese subjects,^52^ only two (*ROS1* and *TP63*) overlapped with ours. Further studies are needed to validate our putative genes in diverse populations and identify ancestry-specific potential causal gene candidates. Second, TWAS methods primarily focus on genes with moderate-to-high *cis*-heritability and high expression levels. Therefore, some genomic regions and low-expression genes were not evaluated. Gene expression prediction performance may be affected by heterogeneities across datasets. Smoking-adjustment analysis cannot rule out all residual confounding effects of cigarette smoking. The EWCE algorithm provides supportive evidence for the identification of cell types where TWAS-implicated genes are enriched instead of causal risk cell types. Finally, small sample sizes of lung tissue ct-eQTL led to the underpowered colocalization. Despite these limitations, our findings provide novel insights into how genetic variants and cell-type–specific expression drive lung carcinogenesis.

In conclusion, our large-scale TWAS identified 109 unique genes associated with risk for overall lung cancer or its histological subtypes. Notably, a subset of risk genes remained significant after adjusting for cigarette smoking, implicating their role in lung cancer biology independent of smoking. Further, in silico analysis and/or *in vitro* experiments prioritized high-confidence causal genes and identified enrichment in specific alveolar and immune cell types. In addition, several genes are targets existing drugs with a potential of reproposing for prevention and clinical treatment. These findings provided valuable insights into the genetic and cellular architecture of lung carcinogenesis, offering new avenues for prevention and treatment.

## Method

### RNA extraction and sequencing

The tumor-adjacent normal lung tissue samples were obtained prospectively from the VTB. After resection, these lung biospecimens were collected and frozen in liquid nitrogen within 2 hours of the resection and then stored at -80°C. Total RNA, including microRNA, was extracted and purified using Qiagen’s AllPrep DNA/RNA/miRNA Universal Kit following the manufacturer’s instructions. The quantity and quality of the RNA samples were checked by Nanodrop (E260/E280 and E260/E230 ratio) and by separation on an Agilent BioAnalyzer. Pair-end whole-transcriptome sequencing was performed using DNBseq. At least 30M reads with a pair-end read length of 100 base pairs were obtained for each sample.

### Gene expression calling and processing

We followed the mRNA analysis pipeline of the GTEx project^54^ and our previous work for RNA-Seq data processing and quality control.^55^ Briefly, the raw read count alignment to the human reference genome version 38 was performed using a two-pass method of the Spliced Transcripts Alignment to a Reference software. Transcript annotations in the human genome were based on GENCODE version 39. Gene-level expressions were quantified from the aligned BAM files using RNAseQC. Gene-level read counts and transcript per million (TPM) values were generated by applying the following read-level filters: 1) reads were uniquely mapped (with a mapping quality of 255 for STAR BAMs); 2) reads were aligned in proper pairs; 3) the read alignment distance was ≤ 6; and 4) reads were fully contained within exon boundaries. Reads that overlapped with introns were not included in the counts. Genes with expression thresholds of > 0.1 transcripts per million (TPM) and ≥ 6 count reads in at least 20% of samples were retained for the downstream analysis. We applied log_2_(TPM+1) transformation, quantile normalization across samples, and then inverse normalization on each gene. Probabilistic estimation of expression residual (PEER) factors^56^ were calculated as GTEx suggests. To account for the potential confounding effects, residuals were calculated by regressing normalized protein levels on age (continuous), biological sex (male or female), cigarette smoking status (ever or never), top three genetic principal components (PCs), and 45 PEERs in multivariate linear regression models. We further conducted rank-based inverse normalization on them to ensure the residuals were normally distributed.

Processed lung tissue gene expression and genotype data from 466 individuals of European ancestry available in the GTEx v10 were used. To obtain the residuals, we adjusted for the confounding effects of age, sex, smoking status, whole genome sequencing (WGS) platform (HiSeq 2000 or HiSeq X), WGS library construction protocol (PCR-based or PCR-free), top three genetic PCs, and 60 PEER factors. We further conducted rank-based inverse normalization on them to ensure the residuals were normally distributed.

### Genotype quality control

Lung tissue DNA samples were genotyped using BioVU MEGA array at Vanderbilt University Medical Center. We followed our previous pipeline to perform quality control^55^. For the following QTL analysis, we kept SNPs with high imputation quality scores (R^2^ > 0.3) and minor allele frequency (MAF) > 5%. For the following prediction model building, SNPs with high imputation quality scores (R^2^ > 0.8), MAF > 5%, and available in GWAS meta-analysis summary statistics were retained. Allele frequencies of palindromic variants were compared with individuals of European ancestry from the 1000 Genome Project (1000GP). No palindromic variants with huge allele frequency differences (> 20%) between the two datasets were identified. WGS data from the GTEx participants was processed in the same way as our previous research.^55^

### GWAS meta-analysis

For meta-analysis, we used lung cancer GWAS summary statistics of a published GWAS meta-analysis of lung cancer^31^ and publicly available GWAS summary statistics from three large biobanks, including the UK Biobank,^57^ FinnGen release version 12,^58^ and the Million Veteran Project.^59^ All these summary statistics were from European descendants. We only kept one common variant for duplicated genetic variants and excluded rare variants with MAF < 1%. Allele frequencies of palindromic variants were compared with those of individuals of European ancestry from 1000GP. Those with MAF of > 40% and an allele frequency difference of > 20% were further removed. We used *METAL* software to combine results from the above studies using the fixed-effect inverse variance weighted method.^60^ Additionally, we performed GWAS meta-analysis of the three main histological subtypes: LUAD, LUSC, and SCLC. The sample size of each GWAS summary statistic was shown in **Table S1**.

### Genetic prediction model building and association analysis

We followed our previous procedures to build gene expression prediction models.^55,61^ We selected variants available in both 1000GP and the GWAS summary statistics. Variants located within 0.5 Mb downstream and upstream of each gene were selected. The elastic net models (*α* = 0.5) with five-fold cross-validation were constructed to predict the expression level of each gene using R *glmnet* package. Reliable prediction models were defined as Pearson’s R > 0.1 and *P* < 0.05.

Each highly predictive model was applied to the lung cancer GWAS meta-analysis summary statistics using the S-PrediXcan framework^62^ in order to evaluate the associations of genetically proxied gene expression levels with lung cancer risk. The formula is shown below:

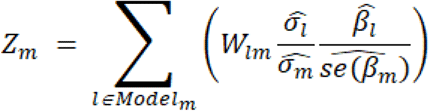

Briefly, a *Z* score was calculated to estimate the direction and strength of associations between the genetically predicted gene expression level and lung cancer risk. W_lm_ represents the weight of genetic variant *l* in the prediction model for a gene *m*. Terms 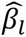 and 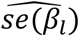 refer to the beta coefficient and the corresponding standard error for genetic variant *l* for lung cancer risk in the GWAS meta-analysis summary statistics, respectively. 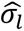 and 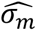 denote the estimated variances of genetic variant *l* and the predicted expression level of gene *m*, respectively.

### Histological subtypes analysis

To identify the histological subtype-specific genes, we performed stratification analysis by three major lung cancer histological subtypes. We applied the highly predictive models to the lung cancer histological subtype-specific GWAS meta-analysis summary statistics using the S-PrediXcan framework.

### TWAS aggregation across the training studies

We then applied the ACAT to combine *P* values of TWAS association results across the VTB and the GTEx for each histological type, respectively. Briefly, the ACAT approach first transforms *P* values to follow a Cauchy distribution under the same null hypothesis and then averages them as the test statistic.

### Conditional analysis of smoking

In order to mitigate residual effects of smoking on lung cancer susceptibility genes, we first performed mtCOJO analyses on GWAS of overall lung cancer risk and its three main histological types conditioned on the largest European-Ancestry GWAS of cigarettes per day.^22,23^ This conditional is more robust to potential collider bias and showed its power to remove the confounding effects of smoking in lung cancer research.^6^ WGS data of 489 Europeans from the 1000GP were used to estimate LD. We then integrated the mtCOJO-corrected GWAS results into genetic prediction models from the two training studies and applied the ACAT to aggregate both studies’ TWAS results.

### Bayesian colocalization between eQTL and GWAS

For those TWAS-identified genes, we conducted colocalization analyses to determine whether gene expression and lung cancer risk share a causal genetic variant. We performed c*is*-eQTL analysis using QTLtools, adjusting for confounders as in our TWAS analysis. Then we meta-analyzed VTB and GTEx c*is*-eQTL summary statistics using METAL.^60^ This analysis was conducted using the R *coloc* package.^19^ We used the default prior probabilities (PP) of P_1_ = P_2_ = 10^−4^ and P_12_ = 10^−5^. The colocalization analysis assessed five hypotheses regarding PP related to potential shared genetic variants between the two traits. A PP of H4 (causal variants shared by both traits) > 0.7 (70%) was defined as strong evidence for sharing a genetic signal between eQTL and lung cancer GWAS.

### Mendelian randomization

SMR was used to identify potentially causal genes associated with lung cancer risk^20^. We used c*is*-eQTL and lung cancer GWAS summary statistics as described above. We set *P* value threshold to select the top associated eQTL as 1×10^-5^ and other default parameters. We used the conservative nominal *P* > 0.05 from the heterogeneity in dependent instruments (HEIDI) test to suggest that the absence of linkage likely influences the main SMR findings. The genes with Bonferroni corrected *P* < 0.05 within each histological subtype and *P*_HEIDI_ > 0.05 were considered as potential causal genes for lung cancer risk.

### Gene set enrichment analysis of TWAS-identified genes

Protein-coding genes identified by our TWAS were included for gene set enrichment analyses to explore the potential molecular pathways for lung cancer risk. The *enrichR* webtool was used for biological function and pathway analyses among *Reactome* 2024. The *Reactome* pathways provide more detailed biological functions than the *KEGG* pathways. Only pathways enriched with more than two genes and an FDR < 5% were considered as significant results.

### Searching drug repurpose targets

To identify potential therapeutic targets, we evaluated proteins encoded by the TWAS-identified genes using three major drug-protein interaction databases, including DrugBank (version 5.1.13)^24^, ChEMBL (version 36, updated on July 28, 2025)^25^, and TTD^26^. DrugBank provides curated information on >500,000 drugs and thousands of associated proteins, genes, and other molecular entities. ChEMBL contains detailed annotations for approximately 2.9 million compounds, 15,600 drugs, and 17,800 targets aggregated from 336 contributing datasets. TTD includes druggability profiles for 426 approved targets, 1,014 clinical trial targets, 212 preclinical or patented targets, and 1,479 targets reported in the literature. From these databases, we extracted drugs and their annotated protein targets and linked them to the proteins encoded by our identified genes. Our mapping focused mainly on drugs approved or under Phase II or III clinical trials.

### Cell type enrichment of TWAS results

We utilized the EWCE algorithm^15^ to assess which cell types the TWAS-identified genes are enriched in. We acquired raw count data of normal lung parenchyma and respiratory airway tissues scRNA-seq data among 63 self-reported Europeans from the Human Lung Cell Atlas.^16^ We used level 3 cell type annotation. We chose overall lung cancer-associated and histological subtype-specific genes significant after Bonferroni correction as the input sets. The bootstrap enrichments calculation was repeated for 10,000 randomly generated gene lists. The cell type-specific enrichment was represented by the standard deviation from the mean computed. *P* < 0.05 was defined as potentially enriched cell types.

### Cell-type-specific eQTL colocalization

We further assessed whether TWAS-identified genes shared the common causal variants with lung cancer risk in each putative risk cell type through Bayesian colocalization. We acquired publicly available lung tissue-based pseudobulk cell type eQTL mapping summary statistics.^65^ Briefly, approximately 66.7% of participants are self-reported Europeans, and the sample size for ct-eQTL ranges from 42 in plasmacytoid DCs to 113 in Monocyte-derived macrophages. We followed the same pipeline using the R *coloc* package^19^ as our bulk tissue eQTL colocalization did. For each gene and cell type, strong evidence of colocalization was defined as a PP of H4 > 0.7.

### Knockout and knockdown experiments in lung cancer cell lines

The latest CRISPR-Cas9 knockout data for the protein-coding genes identified in this study in 126 lung cancer cell lines were acquired from DepMap (version Public 25Q3). The *Chronos* score is a new algorithm for inferring gene fitness effects from CRISPR knockout screens, with lower scores indicating a more essential role of the gene.^18^ Genes with a *Chronos* score of < -0.5 were defined as essential for lung cancer cells proliferation.

We also performed siRNA knockdown experiments in two non-small lung cancer cell lines (A549 and PC-9) for three selected TWAS-identified genes. The human lung cancer cell lines A549 were obtained from the American Type Culture Collection (Manassas, VA, USA) and PC-9 were obtained from Sigma. Cell proliferation, colony formation, immigration, and invasion assays were performed following methods described in previous studies.^63,64^ All *in vitro* functional assays were repeated in three experiments. Differences between experimental groups and controls were assessed using paired t-tests. Significant functions for each gene were defined as nominal *P* < 0.05 in both cell lines.

## Ethics statement

Deidentified data used in this study were derived from published genome-wide association studies or GTEx or public data that followed the relevant institutional review boards and participant consent procedures. This study was approved by Vanderbilt University Institutional Review Board (IRB #202101).

## Data availability

Transcriptomic and genotype data from VTB study are available in the dbGaP database upon manuscript publication. Controlled-access data require approval from the dbGaP Data Access Committee. Requests for access can be submitted through the official dbGaP portal. GENCODE annotation files are available from https://www.gencodegenes.org/human/release_39.html. The publicly available WGS data of GTEx participants used in this study are available in the dbGaP under accession code phs000424.v8.p2. GTEx eQTL summary statistics were downloaded from https://www.gtexportal.org/home/downloads/adult-gtex/qtl. The Transdisciplinary Research in Cancer of the Lung team of the International Lung Cancer Consortium (TRICL-ILCCO) GWAS summary statistics were obtained from dbGaP under accession code phs001273.v1.p1. The processed GWAS summary statistics of UK Biobank, FinnGen R12, and the Million Veteran Project were downloaded from https://finngen.gitbook.io/documentation/data-download. The Human Lung Cell Atlas core scRNA-seq data were downloaded at https://cellxgene.cziscience.com/collections/6f6d381a-7701-4781-935c-db10d30de293. Lung tissue ct-eQTL summary statistics were acquired from https://www.ncbi.nlm.nih.gov/geo/query/acc.cgi?acc=GSE227136. DepMap CRISPR-Cas9 screen experiments data were downloaded from https://depmap.org/portal/. Drugs and chemical compounds data were extracted from publicly available databases, including DrugBank (https://go.drugbank.com/), ChEMBL (https://www.ebi.ac.uk/chembl/), and TTD (https://ttd.idrblab.cn/).

## Code availability

Publicly available software and packages were used throughout this study according to the developer’s instructions. Adopted parameters were specified in this manuscript.

*R version 4.4.0*: https://www.r-project.org/

*R PEER package*: https://github.com/PMBio/peer

*S-PrediXcan*: https://github.com/hakyimlab/MetaXcan

*PLINK version 2*: https://www.cog-genomics.org/plink/2.0/

*TOPMed Imputation Server*: https://imputation.biodatacatalyst.nhlbi.nih.gov/

*METAL*: https://github.com/statgen/METAL

*STAR*: https://github.com/alexdobin/STAR

*RNASeQC*: https://github.com/getzlab/rnaseqc

*QTLtools*: https://github.com/qtltools/qtltools

*SMR*: https://yanglab.westlake.edu.cn/software/smr/

*mtCOJO*: https://yanglab.westlake.edu.cn/software/gcta/#mtCOJO

*R coloc package*: https://github.com/chr1swallace/coloc

*enrichR*: https://maayanlab.cloud/Enrichr/

*Python Scanpy module:* https://github.com/scverse/scanpy

*R EWCE package*: https://github.com/NathanSkene/EWCE

## Acknowledgement

The authors appreciate the contributions and efforts of the researchers and staff of VTB and GTEx. We also thank Ms. Kathleen Harmeyer for assistance with manuscript editing and preparation. This work was conducted in part using the resources of the Advanced Computing Center for Research and Education at Vanderbilt University.

## Author contributions

S.X. J.L. and Q.C. conceived the study and designed the analysis. J. L. and Q.C. supervised the study. S.X., J.S., B.L., R.T., H.C., W.W., Y.Y., X.G., J.L., and Q.C. analyzed data. L.X. performed siRNA knockdown experiments. S.X., J.S., and Q.C. interpreted the data and drafted the original manuscript. All the authors significantly contributed to reviewing and revising the manuscript.

